# Early combination with remdesivir, nirmatrelvir/ritonavir and sotrovimab for the treatment of COVID-19 in immunocompromised hosts

**DOI:** 10.1101/2023.09.07.23295202

**Authors:** Ivan Gentile, Maria Foggia, Maria Silvitelli, Alessia Sardanelli, Letizia Cattaneo, Giulio Viceconte, Federico II COVID team

**Affiliations:** Department of Clinical Medicine and Surgery, Section of Infectious Diseases, University of Naples “Federico II”, Via Sergio Pansini, 5 – 80131, Naples, Italy

**Keywords:** COVID-19, immunocompromised, sotrovimab, remdesivir, nirmatrelvir

## Abstract

**Background:** Immunocompromised patients with COVID-19 have higher morbidity and mortality than general population. Some authors have successfully used antiviral combination, but never in the early phase of the infection.

**Methods:** Retrospective cohort study to describe efficacy and safety of the combination of 2 antivirals, with or without a mAb, both in early (within 10 days from symptoms) and in later phase (after 10 days) of SARS-CoV-2 infection in immunocompromised patients admitted to our facility.

**Results:** We treated 11 patients (7 in early phase and 4 in later phase of COVID-19) with 10 days of intravenous remdesivir plus 5 days of oral nirmatelvir/ritonavir, also combined with sotrovimab in 10/11 cases.

Notably, 100% of the “early” patients reached virological clearance at day 30 from the end of the therapy and were alive and well at follow-up, whereas corresponding figures in the “late” patients were 50% and 75%. Patients in late group more frequently needed oxygen supplementation (p=0.015) and steroid therapy (p=0.045) during admission and reached higher a COVID-19 severity (p=0.017).

**Discussion:** The combination of antiviral and sotrovimab in early phase of COVID-19 in immunocompromised patients is well tolerated and associated with 100% of virological clearance. Patients treated later have lower response rate and higher disease severity, but a causative role of the therapy in such finding is yet to be demonstrated.

## 1. Background

Immunocompromised patients are at high risk of progression and death for COVID-19[1]. Moreover, patients with impaired humoral immunity have the highest risk of prolonged infection, viral and clinical rebound, and the highest mortality rate among immunocompromised patients. [1] Conditions associated the impaired humoral immunity include B-cell depletion due to primary conditions (X-linked agammaglobulinaemia, common variable immunodeficiency, other primary hypogammaglobinaemia) or secondary causes (anti-CD20 treatment in the past year; chronic lymphoblastic leukaemia, non-Hodgkin lymphoma, multiple myeloma).[2]

These patients can develop “persistent inflammatory seronegative COVID”: a novel definition for COVID-19 with prolonged or relapsing symptoms and persistent SARS-CoV-2 RT-PCR on respiratory sample, in absence of serological response to the infection after 14 days. [2]

In these cases, there is no established therapy that can both reduce risk of progression and the time to virological clearance. Although some authors have described the use of antiviral combination with or without monoclonal antibody (mAb), but only in in only in prolonged/relapsing COVID-19 and never in early phase of the infection[3–6]

Since April 2023, we started to use combination with two antivirals with mAb both in prolonged/relapsing COVID-19 and as early treatment (within 10 days from symptoms onset) in immunocompromised patients at high risk of developing prolonged/relapsing infections.

## 2. Methods

Retrospective cohort studies including all the patients with non-critical, mild-to-severe symptomatic COVID-19, admitted to Infectious Disease Ward of Federico II University Hospital from April to July 2023 and treated with at least two antivirals with or without a mAb active o presumably active against the circulating variant at the time of the study period.

The off-label administration of the drugs was approved case-by-case by the Italian regulatory agency on drugs (Agenzia Italiana del Farmaco-AIFA), the Chief Medical Officer of Federico II University Hospital and the Chief of the Hospital Pharmacy, as required by the Italian law and the Hospital’s regulation.

The statistical analysis was performed using SPSS version 27 (SPSS Inc. Chicago, IL). Continuous variables were reported as median and interquartile range and categorical variables as frequencies and percentages. Categorial variables were confronted with Chi-squared test and Fisher’s exact test, when appropriate.

Continuous variables were confronted with Mann-Whitney U test. A significance level of 0.05 was set for the interpretation of the results. Multivariable analysis was not performed due to limited number of events.

## 3. Results

During the observation period we treated 11 immunocompromised with 10 days of intravenous remdesivir plus 5 days of oral N/r, also combined with sotrovimab in 10/11 cases at the fifth day after remdesivir initiation.

Seven of 11 patients received combination therapy within 10 days from COVID-19 symptoms onset (“early treatment group”), while 4/11 after a median of 79 (IQR 48-112) days (“late treatment group”).

All the patients had hematologic malignancies, except one who had Good’s syndrome associated with malignant thymoma and another one who received rituximab for multiple sclerosis. Demographic and clinical characteristics of the patients are shown in **Table 1**.

**Table 1.** Demographic and clinical characteristics. *TIX-CIL: tixagevimab-cilgavimab; N/r: nirmatrelvir/ritonavir; NFS: nasopharyngeal swab; FU: follow-up; WHO: World Health Organization; ADR: adverse drug reaction; CRP: C-reactive protein*.

|  | **Overall**<br>**N=11** | **Early combination**<br>**treatment**<br>**N=7** | **Late combination**<br>**treatment**<br>**N=4** | **p-**<br>**value** |
| --- | --- | --- | --- | --- |
| **Age, years, median (IQR)** | 51 (36-57) | 52 (35-56) | 63 (42-77) | 0.667 |
| **Females, n (%)** | 5 (45.5) | 3 (43) | 2 (50) | 0.652 |
| **Charlson Comorbidity Index, median (IQR)** | 0.5 (0-1.25) | 0 (0-1) | 2 (0.25-3) | 0.517 |
| **Vaccinated, n (%)** | 8 (72) | 5 (71) | 3 (75) | 0.721 |
| **SARS-CoV-2 vaccine doses, median (IQR)** | 3 (3) | 3 (3) | 3 (3) | 1 |
| **Hematologic malignancy** |  |  |  |  |
| Acute myeloid leukemia | 3 (27) | 3 (42) | 0 (0) | 0.3 |
| Non-Hodgkin lymphoma | 4 (36) | 2 (28) | 2 (50) | 0.646 |
| Chronic lymphatic leukemia | 1 (9) | 1 (14) | 0 (0) | 0.308 |
| Multiple myeloma | 1 (9) | 0 (0) | 1 (25) | 0.462 |
| **Other immunodeficiencies, n (%)** | 2 (18) | 1 (14) | 1 (25) | 0.538 |
| **Use of anti-CD20, n (%)** | 2 (18) | 0 (0) | 2 (50) | 0.109 |
| **Deaths, n (%)** | 0 (0) | 0 (0) | 0 (0) |  |
| **Length of stay, days, median (IQR)** | 14 (12.5-23) | 13 (11.5-16.5) | 21 (12.5-38) | 0.2 |
| **Early COVID-19 therapy, n (%)** | 1 (9) | 0 (0) | 1 (25) | 0.364 |
| **Preventive TIX-CIL, n (%)** | 1 (9) | 1 (14) | 0 (0) | 0.325 |
| **Previous therapy for COVID-19, n (%)** | 3 (27) | 0 (0) | 3 (75) | 0.8 |
| **Days from COVID-19 symptoms to combination therapy, median (IQR)** | 9 (3.75-48) | 5 (3-9) | 79 (48-112) | 0.017 |
| **Remdesivir 10 d + N/r, n (%)** | 11 | 7 | 4 |  |
| **Remdesivir 10 d + N/r + Sotrovimab, n (%)** | 10 | 6 | 4 |  |
| **SARS-CoV-2 infection duration from the end of therapy, days, median (IQR)** | 10 (7-22.5) | 11 (8-28) | 8 (3.75-16) | 0.183 |
| **Negative NFS at 14d, n (%)** | 4 (36.4) | 4 (57) | 0 (0) | 0.106 |
| **Negative NFS at 30d, n (%)** | 9 (81) | 7 (100) | 2 (50) | 0.109 |
| **alive and well at FU, n (%)** | 10 (90) | 7 (100) | 3 (75) | 0.364 |
| **Days of follow up, median (IQR)** | 44 (10-92) | 40 (1-81) | 65 (11-108) | 0.183 |
| **Use of immunomodulatory drug for COVID-19, n (%)** | 2 (18) | 0 (0) | 2 (50) | 0.109 |
| **COVID-19 steroid treatment, n (%)** | 6 (54) | 2 (29) | 4 (100) | 0.045 |
| **Worst WHO severity grade, median (IQR)** | 3 (3-4.2) | 3 (3) | 4.5 (4-5) | 0.017 |
| **Need for O2, n (%)** | 5 (45.5) | 1 (14) | 4 (100) | 0.015 |
| **ADR, n (%)** | 1 (9) | 1 (14) | 0 | 0.636 |
| **Lowest lymphocyte value, cells/mm<sup>3</sup>, median (IQR)** | 0.39 (0.15-0.9) | 0.68 (0.2-1.2) | 0.16 (0.07-0.32) | 0.067 |
| **Highest CRP value, mg/dL, median (IQR)** | 3.8 (0.3-15) | 2.3 (0.3-5) | 17 (8-23.4) | 0.117 |
| **Highest LDH value, IU/mL, median (IQR)** | 287 (231-446) | 251 (225-436) | 488 (297-708) | 0.267 |
| **Highest ferritin value, median (IQR)** | 495 (302-1635) | 424 (253-907) | 2000 (542-2000) | 0.286 |
| **Highest D-dimer, median (IQR)** | 863 (571-1580) | 620 (571-1658) | 1422 (1084-1931) | 0.383 |

Only 1 patient in the late treatment group had received an early therapy for COVID-19 in the first 7 days of symptoms: 3 days course of remdesivir after 2 days from the onset of COVID-19 symptoms, 30 days before receiving the combination therapy. Conversely, three patients in the late treatment group with relapsing COVID-19 had received therapy with intravenous remdesivir for 10 days a median of 32 days (IQR 32-50) before receiving the combination therapy.

Only one patient (in the early group) had and an adverse drug reaction (symptomatic bradycardia), that led to remdesivir discontinuation after 8 days of therapy.

As shown in **Table 1**, the two groups of patients did not differ for baseline demographic and clinical conditions. Notably, 100% of the “early” patients reached virological clearance at day 30 from the end of the therapy and were alive and well at follow-up, whereas corresponding figures in the “late” patients were 50% and 75%, with one patients of this group who died with persistence of infection at the follow-up.

It is noteworthy that patients in late group more frequently needed oxygen supplementation (p=0.015) and steroid therapy (p=0.045) during admission and reached a higher COVID-19 severity according to WHO grading (p=0.017)[7], compared to early treated ones, while no significant differences were found in term of laboratory values associated with COVID severity (lymphocyte count, C-reactive protein, LDH).

## 4. Discussion

Despite previous studies have already reported that the antiviral combination therapy is well tolerated and, in many cases, effective in the treatment of prolonged/relapsing COVID-19, we report the first systematic utilization of such strategy also in the early phase of SARS-CoV-2 infection.

According to our results, the use of early combination treatment with 2 antivirals targeting different viral proteins, together with the monoclonal antibody sotrovimab in the early phase of COVID-19 in immunocompromised patients is effective and well tolerated, with 100% clinical and virological response, compared to a lower response rate in patients treated in a later phase, when COVID-19 have already taken a prolonged/relapsing course.

To our best knowledge, our study is the first to evaluate the efficacy of an early antiviral combination treatment in patients at risk of developing prolonged/relapsing form of COVID-19. Other studies, in fact, have assessed similar combinations, but in patients with an established diagnosis of persistent SARS-CoV-2 infection. For example, Mikulska and colleagues reported a case series of 22 immunocompromised COVID-19 patients, of which 18 received full combination of 2 antivirals (remdesivir plus nirmatrelvir/ritonavir or molnupiravir) and one monoclonal antibody (mAb) and 4 received 2 antivirals only after a median time of 42 (IQR 29–100) days from SARS-Cov-2 infection.[6] The authors demonstrate a response rate at day 14, day 30, and last follow-up of 75%, 73%, and 82%, respectively, with a significantly higher response in patients receiving mAbs.[6]

Conversely, Marangoni and colleagues recently described a case of clinical, radiological, and microbiological success in an immunosuppressed patient treated with a combination of nirmatrelvir/ritonavir (N/r) plus molnupiravir for prolonged symptomatic infection with SARS-CoV-2.[8]

It is still unclear whether the fact that patients in late group had higher COVID-19 severity and need for oxygen and steroids is related to the delay in therapy administration, or to factors that are independent from it.

Nonetheless, the results of our study show that the use of early combination treatment with 2 antivirals targeting different viral proteins together with sotrovimab may cause a complete and long-lasting viral clearance and thus prevent the development of severe disease (need or increase of oxygen supplementation) in patients at risk of developing persistent COVID-19. Studies aimed to compare traditional early single drug therapy with an early combination regimen with different durations of treatment in patients with COVID-19 and impairment of humoral immunity are urgently required, but the results of our findings are encouraging.

## Data Availability

All data produced in the present study are available upon reasonable request to the authors

## Acknowledgment

The Federico II COVID-team: Francesco Antimo Alfè, Luigi Ametrano, Francesco Beguinot, Anna Borrelli, Antonio Riccardo Buonomo, Ferdinando Calabria, Giuseppe Castaldo, Letizia Cattaneo, Luca Cianfrano, Diego Coppola, Mariarosaria Cotugno, Federica Cuccurullo, Alessia d’Agostino, Dario Diana, Francesco di Brizzi, Giovanni Di Filippo, Isabella Di Filippo, Antonio Di Fusco, Federico di Panni, Gaia di Troia, Nunzia Esposito, Mariarosaria Faiella, Nicola Ferrara, Lidia Festa, Maria Foggia, Maria Elisabetta Forte, Ludovica Fusco, Antonella Gallicchio, Ivan Gentile, Antonia Gesmundo, Agnese Giaccone, Carmela Iervolino, Irene Iorio, Antonio Iuliano, Amedeo Lanzardo, Federica Licciardi, Giuseppe Longo, Matteo Lorito, Simona Mercinelli, Fulvio Minervini, Giuseppina Muto, Mariano Nobile, Biagio Pinchera, Daria Pietrolouongo. Giuseppe Portella, Laura Reynaud, Alessia Sardanelli, Marina Sarno, Nicola Schiano Moriello, Maria Michela Scirocco, Fabrizio Scordino, Riccardo Scotto, Maria Silvitelli, Stefano Mario Susini, Anastasia Tanzillo, Grazia Tosone, Maria Triassi, Emilia Trucillo, Annapaola Truono, Ilaria Vecchietti, Giulio Viceconte, Riccardo Villari, Emanuela Zappulo, and Giulia Zumbo.

## Author contributions

G.V., I.G. and M.F. conceptualized this correspondence and coordinated and supervised data collection. M.S., L.C. and A.S. collected data, G.V. and M.S. drafted the initial manuscript, and I.G. and M.F. critically revised it. All authors approved the final manuscript as submitted and agree to be accountable for all aspects of the work.

## Institutional Review Board Statement

All procedures performed in studies involving human participants were in accordance with the ethical standards of the institutional and/or national research committee and with the 1964 Helsinki declaration and its later amendments or comparable ethical standards. The study was approved by the Ethical Committee of the University of Naples Federico II (protocol n. 153/22).

## Informed Consent Statement

Patient consent was waived because the research involves materials (data, documents, records, or specimens) that have been collected solely for non-research purposes (medical treatment or diagnosis). The data are recorded by the investigators in an anonymous manner such that subjects cannot be identified directly or through identifiers linked to the subject.

